# Preprocedural CT-guided anatomical assessment shortens intubation time and improves patient and operator experience during unsedated colonoscopy: a randomised controlled trial

**DOI:** 10.64898/2026.09.22.26363660

**Authors:** Kun He, Ke Pang, Jingjuan Liu, Qipu Wang, Yiyang Min, You Wang, Yuhao Liu, Yaowen Hu, Bowen Tian, Hongwei Wang, Rutong Li, Qingwei Jiang, Qiang Wang, Tao Xu, Shengyu Zhang, Wei Liu, Dong Wu

## Abstract

Looping is a major cause of difficult and painful insertion during unsedated colonoscopy, yet strategies are largely guided by trial and error; prolonged insertion may jeopardise completion, discourage future surveillance and is associated with lower adenoma detection. In this randomised trial of 141 patients, protocolised review of pre-existing abdominal-pelvic computed tomography (CT) anticipated colonic anatomy and shortened caecal intubation time by 136 s, while reducing pain, discomfort and operator workload and increasing willingness to repeat colonoscopy. Effects were largest in N-loop and alpha-loop configurations, which accounted for 83% of patients. Routinely acquired CT could support anatomy-informed colonoscope insertion especially when anesthesia services are limited or sedation poses an elevated risk.

## IN MORE DETAIL

Looping, particularly in the sigmoid colon, is a major determinant of difficult and painful colonoscope insertion. In unsedated or minimally sedated colonoscopy, pain during difficult insertion may prevent completion and discourage future surveillance[1–4]. Prolonged caecal intubation increases distension and procedural burden and has been associated with lower adenoma and advanced adenoma detection[5,6].For operators, difficult loop reduction converts insertion into repeated trial-and-error decisions - withdrawal, torque, abdominal compression and repositioning - increasing cognitive and physical workload[7]. Water-aided colonoscopy[8], caps[9], magnetic endoscope imaging[10] and overtube systems[11] can assist during insertion, whereas CT offers a different opportunity: anticipating patient-specific anatomy before the colonoscope enters the patient. CT reconstruction studies have linked failed colonoscopy to longer colonic segments and more angulations[12], but no randomised trial has tested protocolised preprocedural CT guidance during routine colonoscopy.

We enrolled adults aged 18–75 years undergoing unsedated outpatient colonoscopy with topical oxybuprocaine at Peking Union Medical College Hospital between May and August 2026 **(Supplementary Figure 1)**. Eligible patients had an evaluable abdominal-pelvic CT examination within the preceding 3 years and had undergone no subsequent abdominal or pelvic surgery. The interval was supported by a prestudy validation: among 1000 patients were randomly sampled from approximately 5000 with two CT examinations within 3 years and no intervening surgery, colonic configuration was unchanged in 999 (The only observed change occurred in a patient with substantial worsening of chronic constipation between examinations). All procedures were performed by three endoscopists, each with experience of approximately 3000–5000 colonoscopies.

Patients randomized 1:1 to protocolised CT-guided or conventional insertion. In the CT-guided group, endoscopists traced the colon from rectum to caecum on CT, classified sigmoid anatomy into six predefined configurations (**Figure 1**), documented sigmoid and transverse-colon redundancy, and used this assessment to guide loop reduction and abdominal pressure. When abdominal pressure was required, it was applied over the abdominal wall directly overlying the most cranial extent of the sigmoid colon identified on CT. Controls underwent usual insertion without protocolised CT review, with abdominal pressure applied to the standard infraumbilical site. Withdrawal time was standardized as far as practicable to approximately 6 min in both groups. All colonoscopies in both study groups were performed by three endoscopists, each with experience of approximately 3000–5000 colonoscopies.

**Figure 1.**
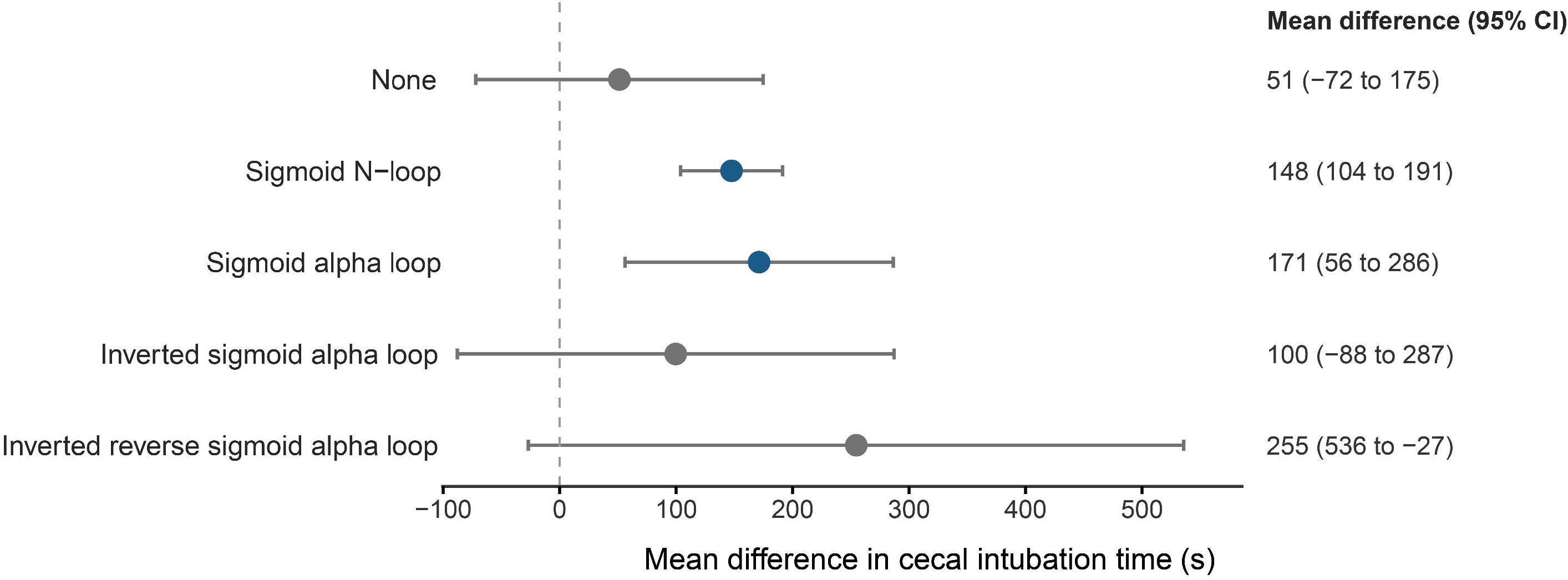
Six predefined sigmoid colon configurations used for CT-based anatomical assessment. (A) No-loop; (B) N-loop; (C) alpha-loop; (D) reverse sigmoid alpha-loop; (E) inverted sigmoid alpha-loop; and (F) inverted reverse sigmoid alpha-loop.

Before recruitment, the three endoscopists completed standardised training with 80 CT cases and correctly classified 10 consecutive test cases. CT review was undertaken during routine preparation and required a median of 5.0 min (IQR [2.1-8.5]). A blinded radiologist independently classified sigmoid configuration (κ=0.86, 95% CI 0.82 to 0.95). Agreement, rather than accuracy against colonoscopy, was assessed because colonoscopy does not visualise shaft configuration. **Online supplemental material S2** presents matched CT and schematic views of the six patterns.

Of 210 patients assessed, 141 were randomised (**Table 1**). CT guidance shortened adjusted caecal intubation time from 391 to 255 s (mean difference -136 s, 95% CI -156 to -116). Patients in the CT-guided group reported less pain, greater willingness to repeat colonoscopy and lower overall discomfort, while operators reported substantially lower workload (**Table 2**). De-looping attempts, abdominal compression attempts and polyp count were similar between groups. The effect was most evident in N-loop and alpha-loop configurations (**Figure 2**).

**Table 1.** Baseline demographic, clinical and CT-defined anatomical characteristics of randomised patients.

| Characteristic | Conventional (n=70) | CT-guided (n=71) |
| --- | --- | --- |
| **Age, years** | 64 ± 11 | 66 ± 9 |
| **Sex** |  |  |
| Female | 29 (41%) | 33 (46%) |
| Male | 41 (59%) | 38 (54%) |
| **Height, cm** | 167 ± 7 | 164 ± 8 |
| **Weight, kg** | 66 ± 12 | 65 ± 13 |
| **Indication for colonoscopy** |  |  |
| Diagnostic | 40 (57%) | 42 (59%) |
| Screening | 14 (20%) | 12 (17%) |
| Surveillance | 16 (23%) | 17 (24%) |
| **BBPS total score** | 6.27 ± 1.76 | 5.76 ± 0.85 |
| **Adequate bowel preparation** |  |  |
| Inadequate | 20 (29%) | 23 (32%) |
| Adequate | 50 (71%) | 48 (68%) |
| **Sigmoid configuration** |  |  |
| No loop | 5 (7.1%) | 8 (11%) |
| **Common sigmoid loop types** |  |  |
| Sigmoid N-loop | 51 (73%) | 51 (72%) |
| Sigmoid alpha loop | 6 (8.6%) | 9 (13%) |
| **Rare sigmoid loop types** |  |  |
| Inverted reverse sigmoid alpha loop | 2 (2.9%) | 1 (1.4%) |
| Inverted sigmoid alpha loop | 5 (7.1%) | 2 (2.8%) |
| Reverse sigmoid alpha loop | 1 (1.4%) | 0 (0%) |
| **Transverse redundancy** | 5 (7.1%) | 6 (8.5%) |
| **Sigmoid redundancy** | 13 (18.6%) | 12 (17.1%) |

Data are mean ± SD or n (%), unless otherwise stated. BBPS, Boston Bowel Preparation Scale.

**Table 2.** Caecal intubation time, patient-reported outcomes and operator workload.

| Domain | Outcome | Conventional (n=70) | CT-guided (n=71) | Effect estimate (95% CI) | p value |
| --- | --- | --- | --- | --- | --- |
| **Caecal** | Caecal intubation | 399 ± 213 | 247 ± 154 | -153 (-214 to -91)<sup>a</sup> | <0.001 |
| **intubation time** | time, s |  |  |  |  |
|  | Model-adjusted mean caecal intubation time, s | 391 (364-418) | 255 (228-281) | -136 (-156 to -116)<sup>b</sup> | <0.001 |
| **Patient experience** | Pain score, 0-10 | 4.3 ± 1.6 | 2.5 ± 1.5 | -1.74 (-2.26 to -1.21)<sup>a</sup> | <0.001 |
|  | Willingness to repeat, 0-10 | 7.0 ± 1.4 | 8.0 ± 1.4 | 1.03 (0.56 to 1.50)<sup>a</sup> | <0.001 |
|  | Overall discomfort, 1-5 | 3 (3-4) | 2 (1-2) | 0.04 (0.01 to 0.08)<sup>c</sup> | <0.001 |
| **Operator workload** | NASA-TLX total score | 49.5 ± 10.3 | 31.1 ± 9.6 | -18.5 (-21.8 to -15.2)<sup>a</sup> | <0.001 |
|  | Mental demand | 49.4 ± 10.5 | 29.1 ± 10.6 | -20.3 (-23.8 to -16.7)<sup>a</sup> | <0.001 |
|  | Physical demand | 50.4 ± 11.4 | 32.3 ± 9.7 | -18.2 (-21.7 to -14.7)<sup>a</sup> | <0.001 |
|  | Temporal demand | 50.1 ± 11.4 | 31.8 ± 9.9 | -18.2 (-21.8 to -14.7)<sup>a</sup> | <0.001 |
|  | Effort | 50.6 ± 10.3 | 31.6 ± 10.2 | -19.0 (-22.5 to -15.6)<sup>a</sup> | <0.001 |
|  | Performance burden | 48.6 ± 10.7 | 32.1 ± 10.2 | -16.5 (-20.0 to -13.0)<sup>a</sup> | <0.001 |
|  | Frustration | 48.1 ± 10.5 | 29.4 ± 9.5 | -18.6 (-22.0 to -15.3)<sup>a</sup> | <0.001 |
| **Other outcomes** | De-looping attempts | 1.3 ± 0.5 | 1.2 ± 0.5 | 0.86 (0.64 to 1.16)<sup>d</sup> | 0.318 |
|  | Abdominal-compression attempts | 0.9 ± 0.5 | 0.9 ± 0.4 | 0.97 (0.68 to 1.38)<sup>d</sup> | 0.866 |
|  | Polyp count | 1.0 ± 0.9 | 1.1 ± 1.1 | 1.12 (0.81 to 1.55)<sup>d</sup> | 0.508 |

Data are mean ± SD or median (IQR), unless otherwise stated. <sup>a</sup>Mean difference. <sup>b</sup>Adjusted mean difference from a linear regression model including operator, height, weight, Boston Bowel Preparation Scale score and CT-defined sigmoid configuration. <sup>c</sup>Odds ratio from ordinal logistic regression. <sup>d</sup>Incidence rate ratio from count regression. CI, confidence interval; IRR, incidence rate ratio; NASA-TLX, National Aeronautics and Space Administration Task Load Index; OR, odds ratio.

**Figure 2.**
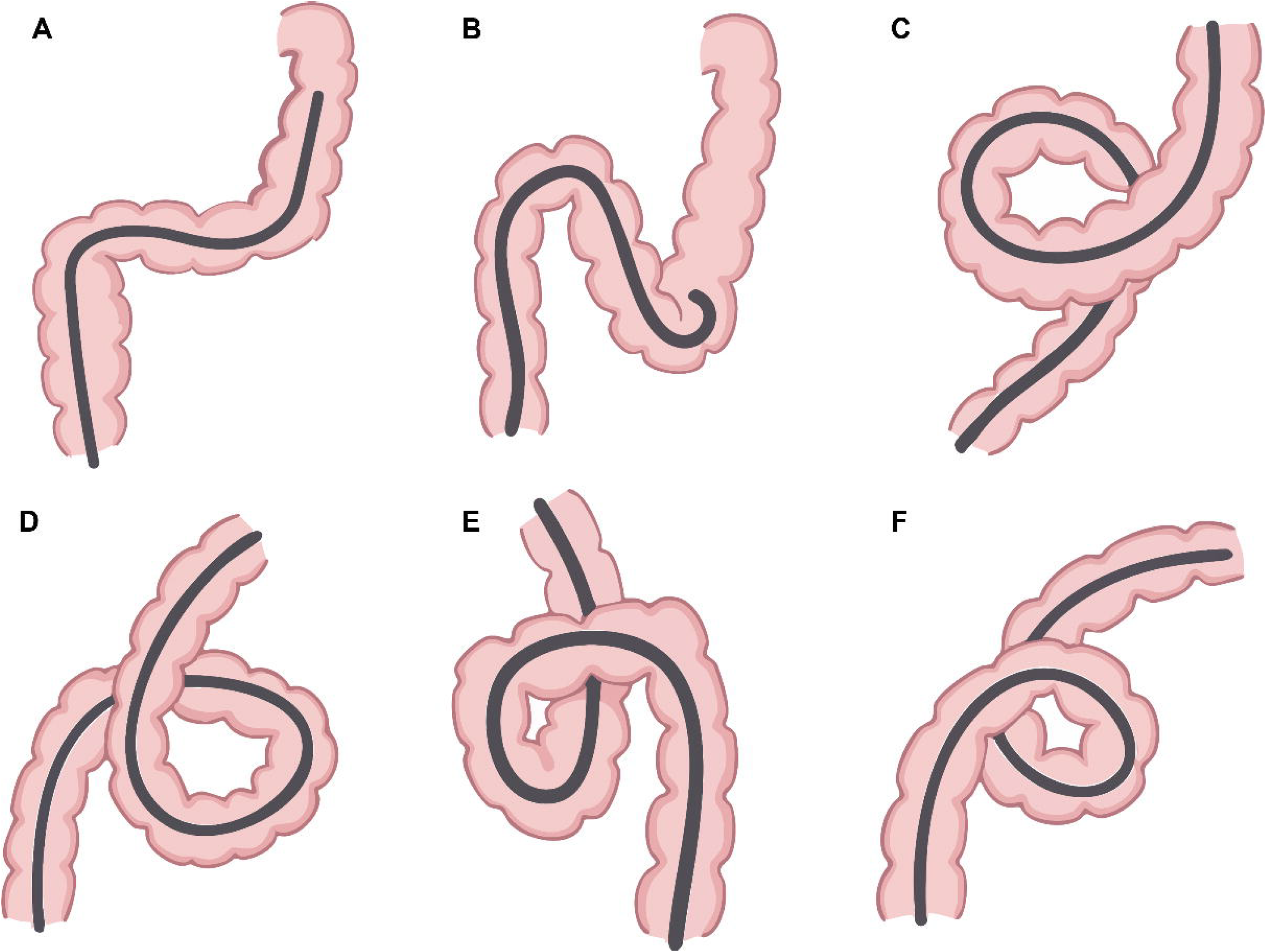
Association between sigmoid loop type and caecal intubation time. Adjusted effects for no loop, N-loop, alpha loop, inverted alpha loop and inverted reverse alpha loop. Estimates are mean differences with 95% CIs; estimates with 95% CIs excluding zero are highlighted in blue.

## DISCUSSION

In this prospective, patient-blinded randomized controlled trial, CT-guided colonoscopy significantly shortened cecal intubation time compared with conventional colonoscopy. In addition to improving insertion efficiency, CT guidance was associated with lower patient-reported pain, greater willingness to undergo repeat colonoscopy, and substantially reduced operator workload across all NASA-TLX domains. Importantly, these benefits were not accompanied by a reduction in polyp count, suggesting that faster insertion did not come at the expense of detection-related performance.

The clinical relevance of these findings is best understood in the context of unsedated or minimally sedated colonoscopy. In appropriately selected patients, it can be an acceptable and practical option that avoids escort requirements, reduces recovery-room use, and allows more rapid return to normal activities[13]. Prior reports have noted that even endoscopists and gastroenterology physicians have chosen scheduled unsedated colonoscopies for their own screening or surveillance[14,15], suggesting that the option can be acceptable when patients are properly informed and procedures are carefully performed. Therefore, improving insertion comfort in unsedated colonoscopy has clinical value beyond pain control alone.

The mechanism is anatomically plausible. In conventional colonoscopy, sigmoid loop formation must be inferred indirectly from resistance, tip movement, patient discomfort and the response to torque, withdrawal, abdominal compression or position change. This reactive process often requires repeated trial-and-error manoeuvres and may partly explain both the prolonged learning curve of colonoscope insertion and the operator workload observed during difficult cases. Randomised evidence from magnetic endoscope imaging supports the value of visualising shaft configuration during insertion[16]. CT guidance offers a preprocedural analogue: it allows the endoscopist to anticipate the likely difficult segment and plan the timing and location of loop reduction, abdominal compression and repositioning before insertion begins. The subgroup findings further support this interpretation. The reduction in caecal intubation time was greatest in patients with N-loop and alpha-loop configurations, which accounted for most patients[10], whereas estimates were smaller or less precise in no-loop and uncommon inverted configurations. Thus, the value of CT guidance may lie less in detecting rare extreme anatomy than in helping endoscopists manage common, recurrent sigmoid loop patterns more deliberately. This interpretation is supported by the reduced NASA-TLX scores despite similar numbers of de-looping and abdominal-compression attempts, suggesting that CT guidance improved the targeting and efficiency rather than the frequency of manoeuvres.

A practical advantage of this strategy is that it uses CT images that were already available before colonoscopy. The present study should not be interpreted as supporting additional CT scanning solely for colonoscopy guidance, given the issues of radiation exposure and cost. Rather, the findings suggest that when recent abdominal-pelvic CT images are already available, they may contain clinically useful anatomical information that is usually underused by endoscopists. This is particularly relevant in patients who undergo CT for diagnostic evaluation, cancer staging, preoperative assessment, or other abdominal conditions before colonoscopy. The ability to extract procedural guidance from existing imaging may offer a low-cost and scalable way to improve colonoscopy performance without introducing new devices or substantial workflow disruption.

Limitations include the single-centre design, the involvement of only three trained endoscopists, inclusion restricted to patients with available CT, and inability to blind operators. Sigmoid loop configuration was the primary anatomical target of the intervention. However, insertion difficulty is also influenced by colonic redundancy, which may require different insertion strategies from those used to manage loop configuration. Accordingly, CT-defined sigmoid and transverse-colon redundancy were recorded at baseline and adjusted for in the primary analyses. Whether CT-guided selection of manoeuvres, such as targeted abdominal compression or patient repositioning, could further facilitate colonoscope insertion in patients with redundant colons remains unknown and deserves investigation in future studies. CT is also static and may not reflect dynamic changes caused by insufflation, position, abdominal pressure or scope manipulation. Patient- and operator-reported outcomes may be susceptible to expectation effects, and subgroup findings should be considered exploratory.

In conclusion, CT-guided anatomical assessment before colonoscopy significantly shortened cecal intubation time, improved patient tolerance, and reduced operator workload without evidence of reduced detection-related performance. When prior abdominal-pelvic CT images are available, CT-guided insertion may provide a practical strategy to convert colonoscope insertion from a reactive trial-and-error process into an anatomically informed procedure. Further multicenter studies are warranted to validate these findings, standardize CT-based assessment, and determine whether this approach can improve broader quality, access, and implementation outcomes in routine colonoscopy practice.

## Supporting information

Sipplementary Material 2

## Data Availability

All data produced in the present study are available upon reasonable request to the authors

## ONLINE SUPPLEMENTAL MATERIAL

**Supplementary Figure S1. CONSORT flow diagram of participant enrolment, allocation, follow-up and analysis**. Flow diagram showing participant screening, randomisation, allocation, follow-up and inclusion in the primary analysis. Of 210 patients assessed for eligibility, 155 were randomised to the computed tomography (CT)-guided group or control group. Seventy-one patients in each group received the allocated intervention. One patient in the control group was excluded from the primary analysis because of a nonstandard sigmoid colon anatomical configuration, resulting in 71 and 70 patients being included in the final analysis, respectively.

**Supplemental Material S2**. Six sigmoid configurations shown in parallel as representative CT images, physical twist-stick models and schematic illustrations: no loop, N-loop, alpha loop, reverse alpha loop, inverted alpha loop and inverted reverse alpha loop.

## Notes

***Funding:*** This study was supported by the Clinical Research Excellence Program for Research-Oriented Wards, Beijing Municipal Health Commission (BRWEP2024W034010103); the National Key Research and Development Program of China (2024YFA0918504); the Beijing Municipal Natural Science Foundation (L232016 and 7232123); the Science and Technology Planning Project of the Tibet Autonomous Region (XZ202501JD0021); National High Level Hospital Clinical Research Funding (2025-PUMCH-C-048); the Chinese Academy of Medical Sciences Innovation Fund for Medical Sciences (2022-I2M-1-003).

### Competing Interest Statement

The authors have declared no competing interest.

### Clinical Trial

NCT07541924

### Author Declarations

The study was approved by the institutional ethics committee of Peking Union Medical College Hospital (Approval Number: I-26PJ1859). All participants provided written informed consent.

