## Supplementary material for "Preprocedural CT-guided anatomical assessment shortens intubation time and improves patient and operator experience during unsedated colonoscopy: a randomised controlled trial": Sipplementary Material 2

### Slide 1
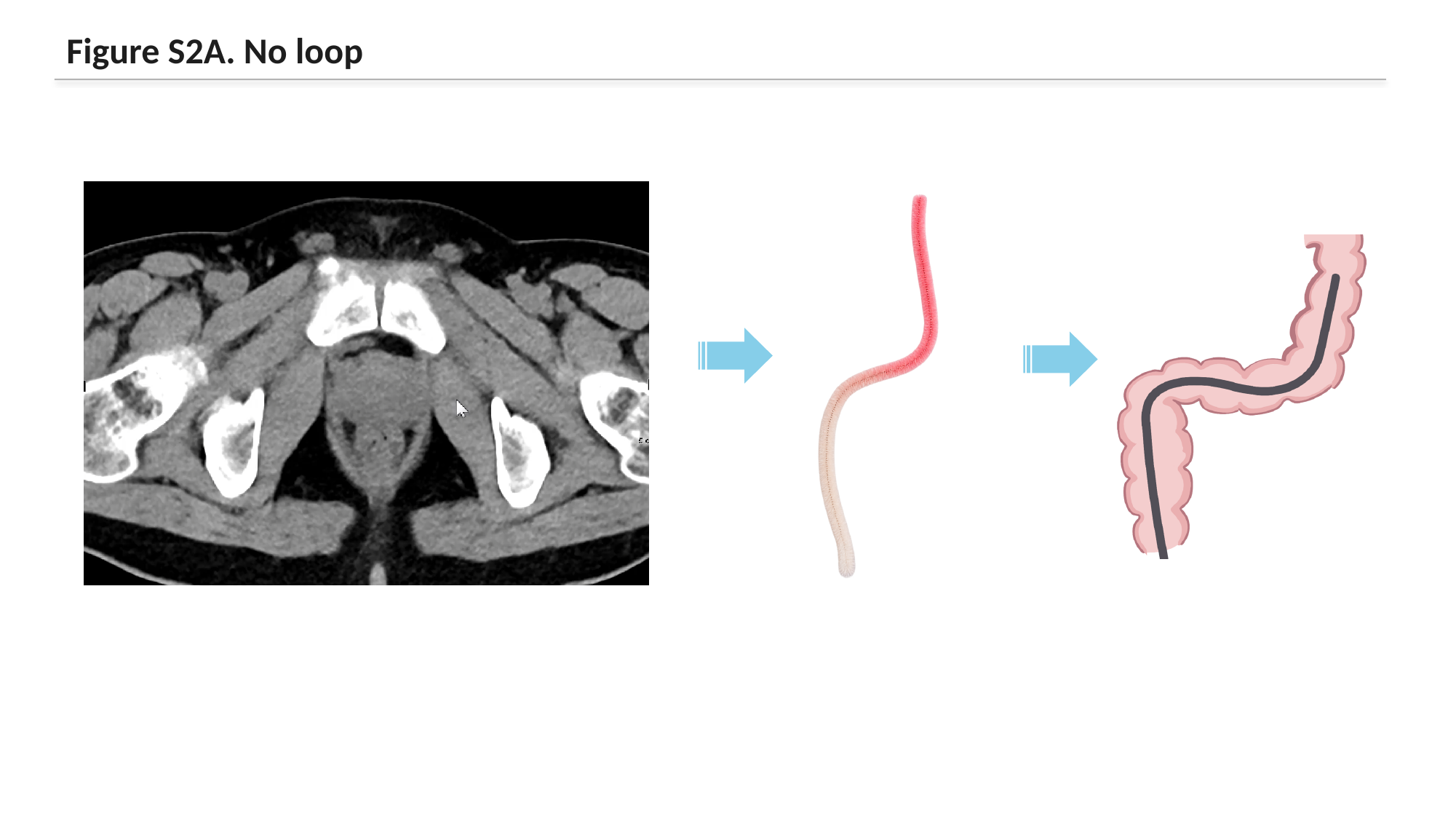

Figure S2A. No loop

### Slide 2
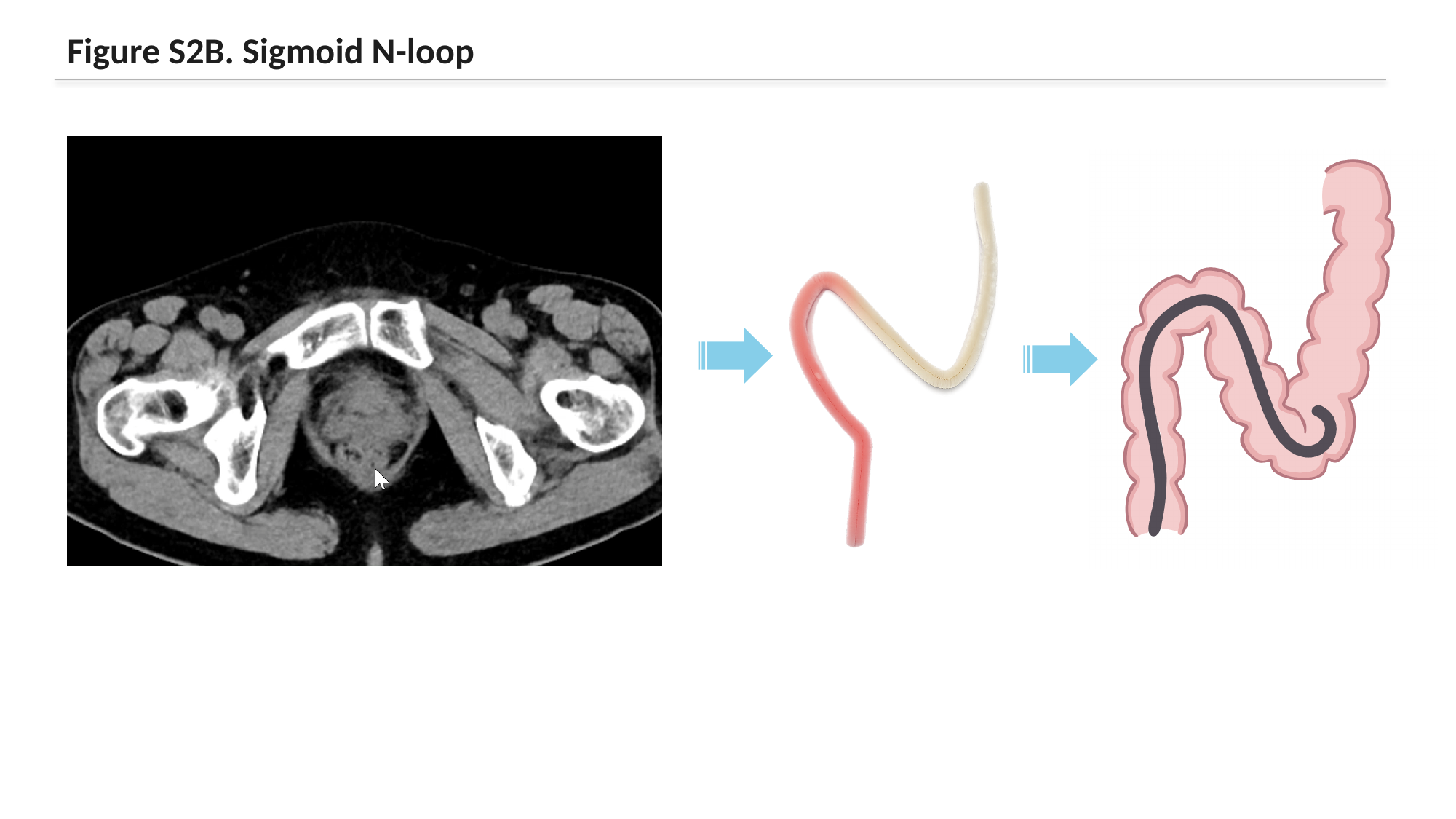

Figure S2B. Sigmoid N-loop

### Slide 3
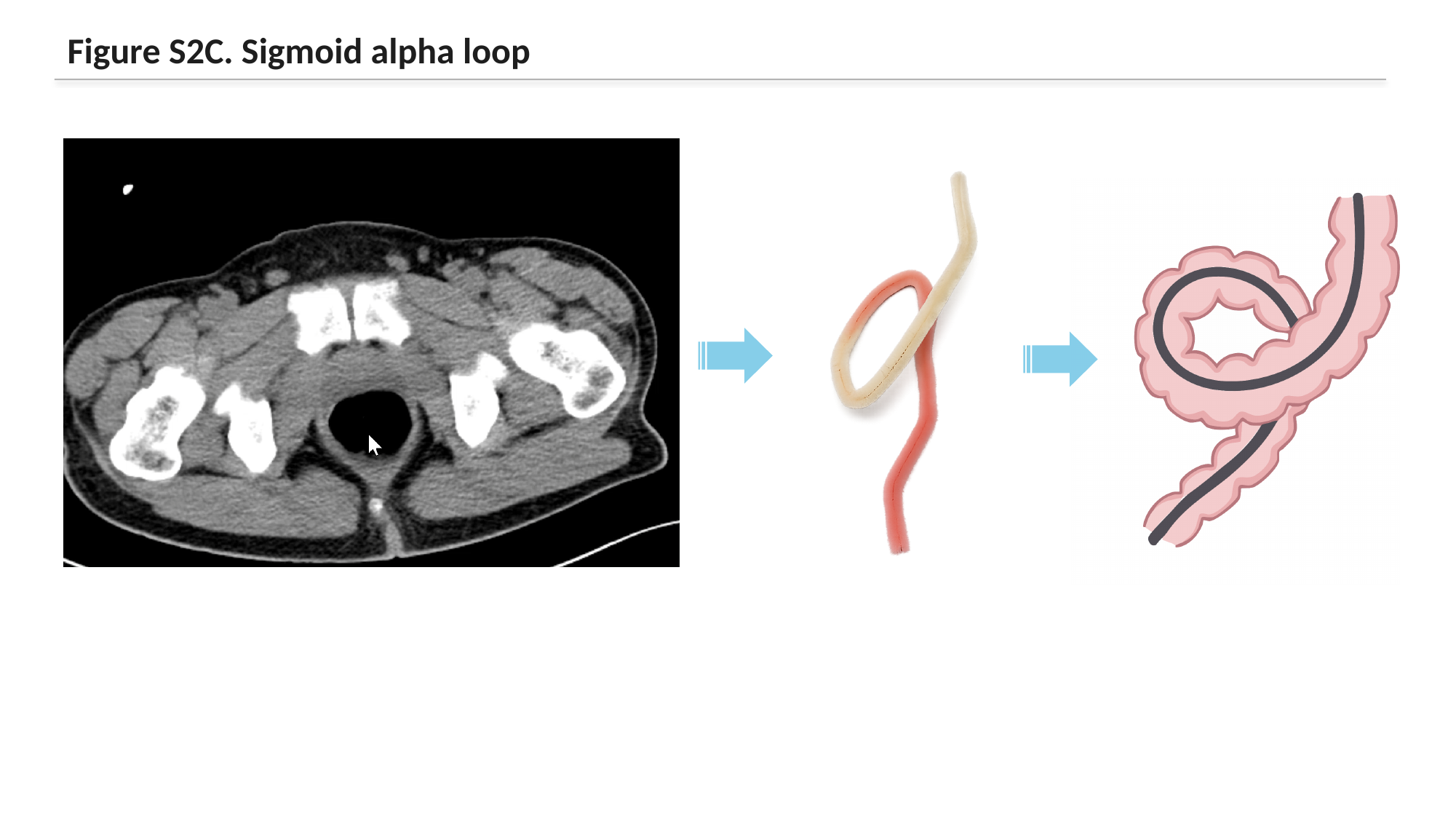

Figure S2C. Sigmoid alpha loop

### Slide 4
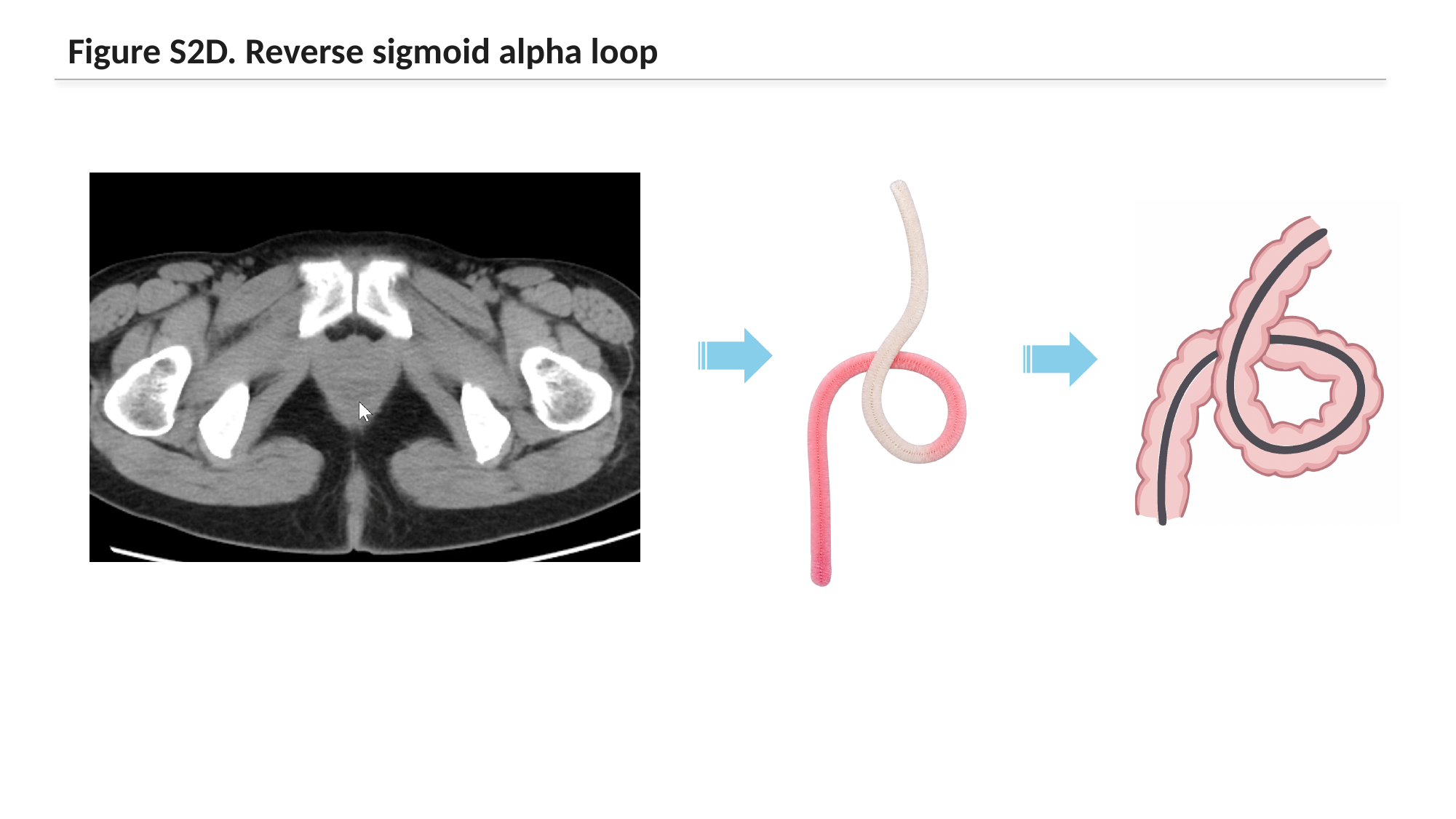

Figure S2D. Reverse sigmoid alpha loop

### Slide 5
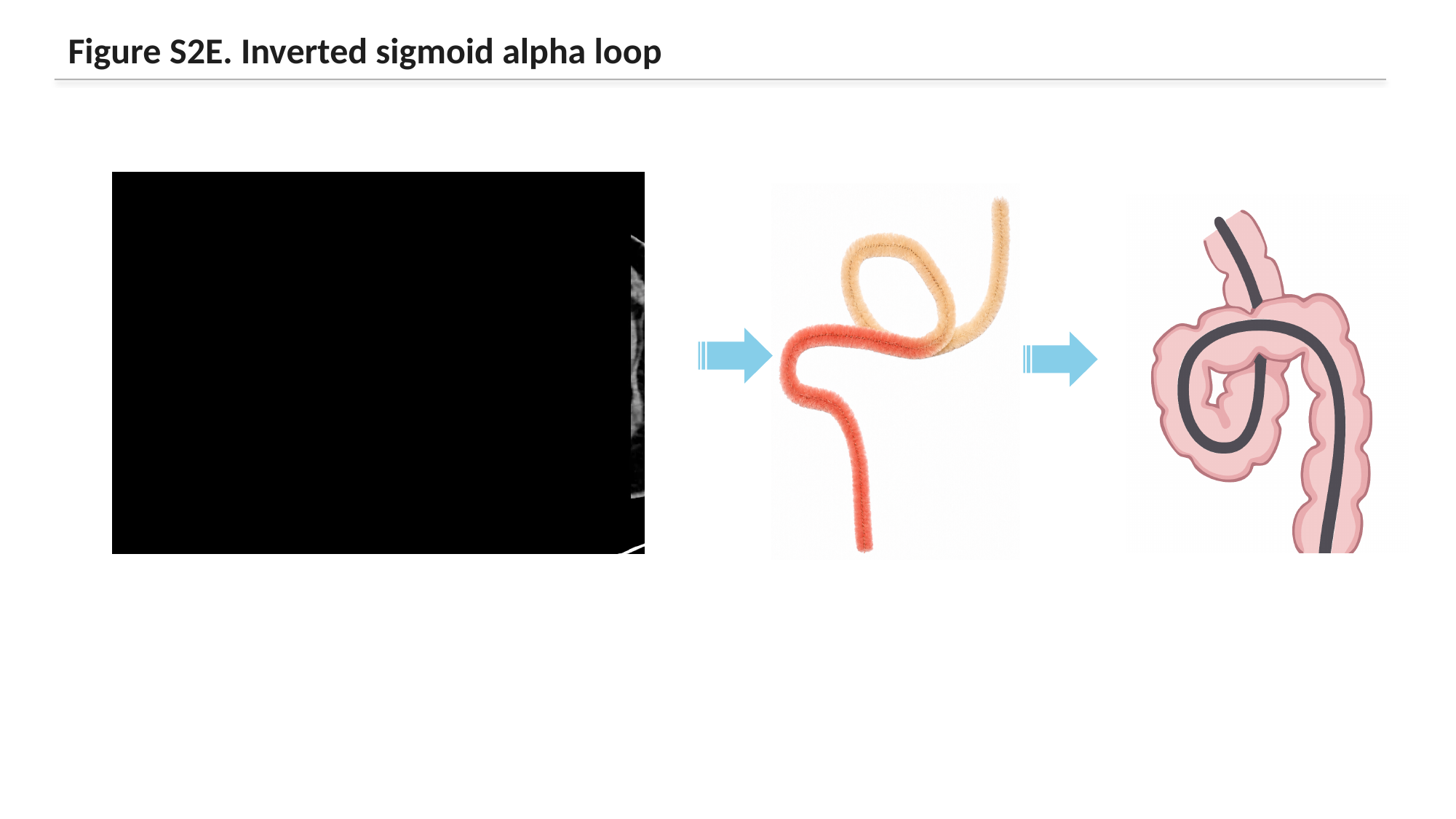

Figure S2E. Inverted sigmoid alpha loop

### Slide 6
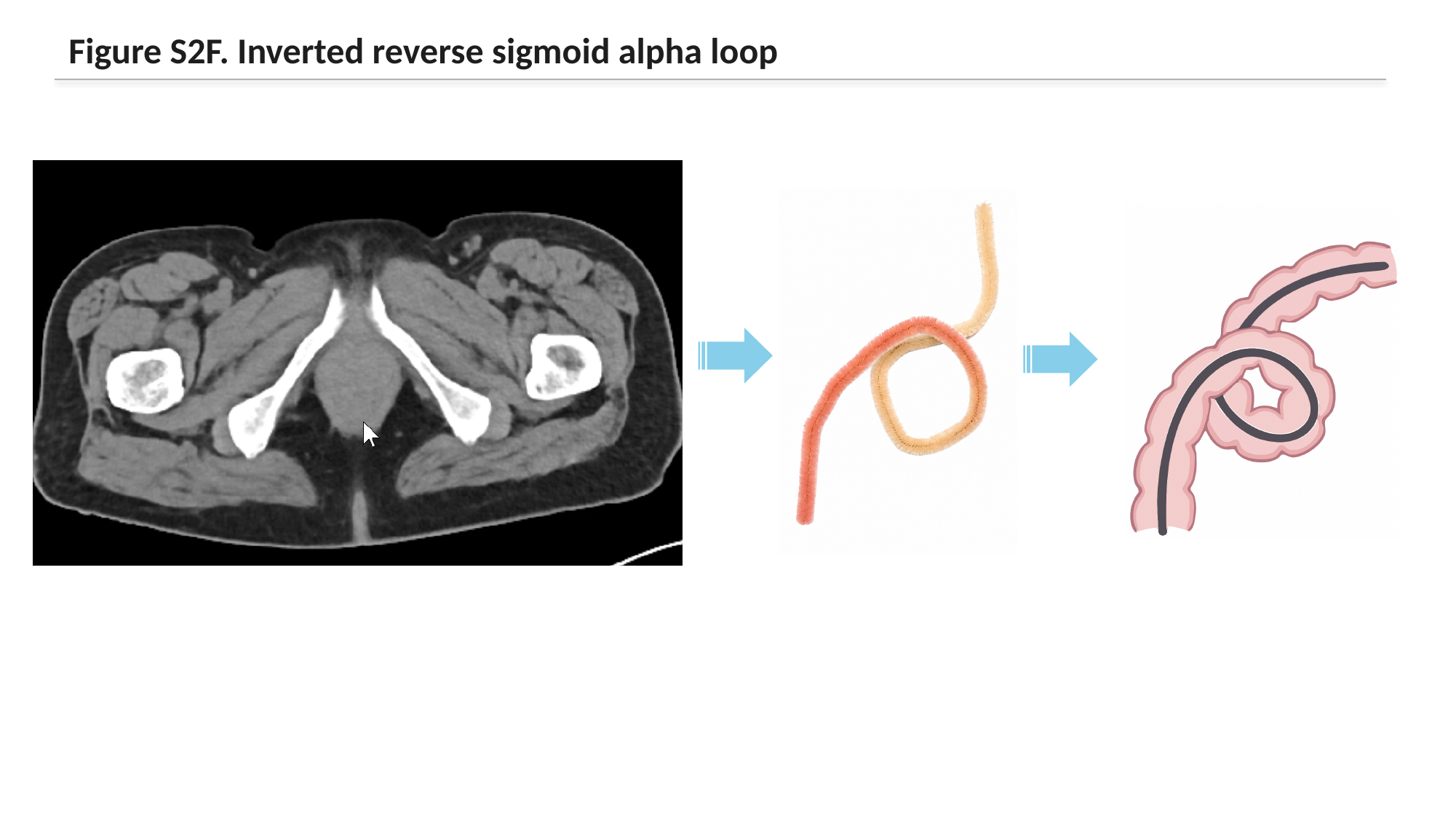

Figure S2F. Inverted reverse sigmoid alpha loop
